# Development of a Rule-Based Knowledge Base for Digitalization of Standard Operating Procedures on Radiotherapy in Breast Cancer

**DOI:** 10.1101/2025.06.05.25329060

**Authors:** Fabio Dennstädt, Johannes Zink, Nikola Cihoric, Dagmar Weder, Markus Glatzer, Julie Crommelinck, Krisztian Süveg, Galina Farina Fischer, Hông-Linh Hà, Detlef Brügge, Ludwig Plassilm, Paul Martin Putora

## Abstract

**Background:** Traditional text-based standard operating procedures (SOPs) present limitations in today’s data-rich environment, being static and difficult to query. We developed a rule-based knowledge base to overcome these challenges through digitalization of SOPs on breast cancer radiotherapy.

**Methods:** We created a web application with a relational database structured around Common Data Elements (CDEs), a rule-based inference engine, and a responsive user interface. The system was tested with nine radiation oncologists over 14 months to evaluate information retrieval success.

**Results:** The knowledge base incorporated 8 main topics, 31 subtopics, 103 information entries, and references to 48 clinical trials and 65 publications. During testing, 56.5% of queries were successful, while analysis of unsuccessful queries revealed most information was available but inaccessible due to user-experience issues.

**Conclusion:** Our rule-based knowledge base demonstrates the feasibility of transforming static SOPs into interactive, evidence-linked resources. While promising, substantial user experience barriers remain. Future enhancements should prioritize user-interface improvements, sustainable knowledge-updating mechanisms, and AI integration to strengthen the system for practical use.

## INTRODUCTION

In modern oncology, clinicians face increasingly complex decision-making scenarios that require comprehensive knowledge of rapidly evolving evidence, classification systems, and molecular characteristics of diseases. For breast cancer specifically, these decisions must account for multiple factors including tumor stage, molecular subtypes, receptor status, patient characteristics, and the continuously expanding body of clinical trial data [1]. The integration of this knowledge into clinical practice presents a considerable challenge, particularly in time-constrained environments.

Traditional approaches to managing medical knowledge in oncology rely heavily on text-based standard operating procedures (SOPs), written guidelines, and individual literature searches.

While these methods have served the medical community for decades, they have inherent limitations in today’s data-rich environment. Information in text documents is typically static, difficult to query for specific contexts, and lacks interconnectivity between related concepts. This creates a substantial burden for clinicians who must compile and integrate information from disparate sources when making treatment decisions.

One approach to overcome this problem is the usage of digitalized guidelines and SOPs with logically connected information inside a knowledge base [2].

Unlike conventional text-based resources, such a system enables precise queries for specific clinical scenarios and provides context-relevant information based on well-defined data architectures. The approach transforms how clinical knowledge is accessed, allowing clinicians to retrieve evidence-based information tailored to various oncological scenarios. Advantages of this approach include enhanced information accessibility and integration, improved decision support, real-time updates and version controls, enhanced collaboration as well as standardization [3] [4]. Furthermore, it allows the computer-based analysis of usage and connection to other software systems, such as electronic healthcare records (EHRs) [5].

In this work, we provide the technical report and experiences of the development of a rule-based knowledge base for digitalization of SOPs on breast cancer in radiation oncology.

## METHODS

### System Requirements

To realize the digitalized SOP on breast cancer, we developed a web application to support medical professionals in reviewing and analyzing clinical guidelines, oncological literature and clinical trials tailored to a given oncological scenario. We defined the following required functionalities for the system:

- Evaluation of recommendations of clinical guidelines: Possibility for the user to navigate through medical criteria, set parameter values, and see how different parameter combinations affect recommendations of the guidelines.
- Access medical trials and publications: Implementation of a database of trials and publications that can be accessed, showing publication details, authors, dates, and relationships between trials. Users can browse through relevant trials and publications filtered by tags and criteria. The application shows which trials are applicable to specific oncological scenarios.
- Rule-based operation: The recommendations shown by the system are made based on explicit formalized rules. These rules can be inspected to understand why a specific recommendation was given.
- Interoperable data system: The criteria and parameters used are clearly defined. Transformation into the concept of Common Data Elements (CDEs) introduced by the National Institutes of Health (NIH) [6] is possible, which ensures consistent and systematic data collection and enables communication with other software systems based on Fast Healthcare Interoperability Resources (FHIR) standards.

A schematic illustration for the conceptualized framework of the rule-based knowledge base is provided in **Figure 1** and will be explained in detail in the following.

**Figure 1:**
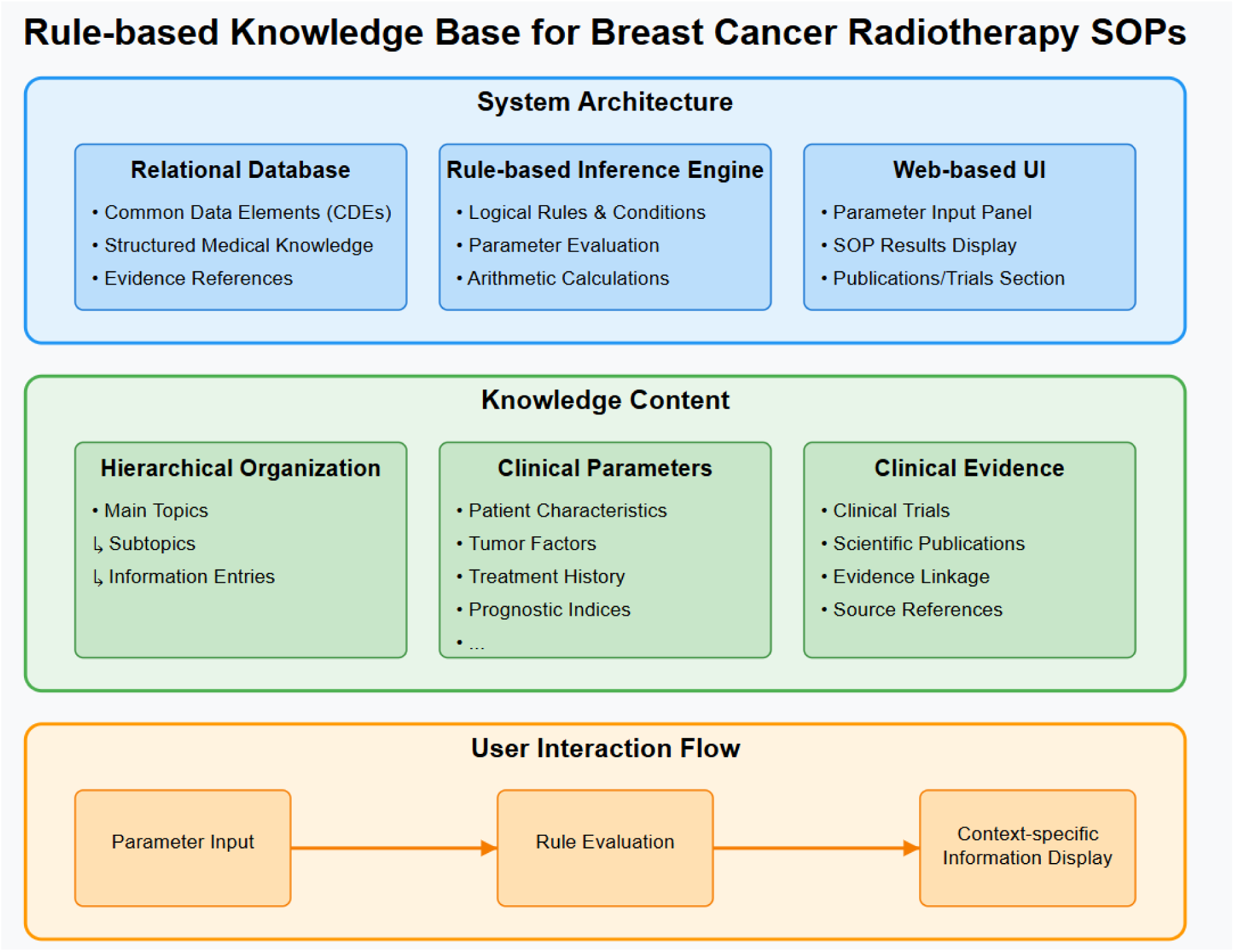
Schematic illustration of the framework for the rule-based knowledge base to digitalize SOPs on breast cancer in radiation therapy.

### System Architecture and Technical Implementation

We designed the knowledge as a structured, interactive data repository for breast cancer radiotherapy information. We implemented a comprehensive data architecture based on CDEs to ensure standardization and interoperability of clinical information. The system comprises three main technical components: (1) a relational database structure, (2) a rule-based inference engine, and (3) a web-based user interface.

### Relational Database, Data Structure and Organization

Based on previous work, we screened the existing text-based SOPs and analyzed for the occurrence of relevant information on which a recommendation is dependent [7]. This information was clearly defined and formalized based on the agreement of two radiation oncologists, who use the SOPs in their daily clinical work as physicians. As a result, corresponding CDEs were defined and the CDE-based data structure was conceptualized as previously published [8]. We created a list of recommendations that are dependent on these CDEs based on the local SOPs. In addition, we defined clinical guidelines as well as publications and trials considered to be relevant for mentioning in the knowledge base on breast cancer. For each entry in the knowledge base (general information, recommendations, publications, trials, guidelines), we defined relevant clinical parameters as CDEs with specific data types (categorical, numerical, or boolean) and permissible value ranges. These parameters serve as both search criteria and contextual filters that determine which information is relevant in specific clinical scenarios. We developed a separate editor system for creating/editing of the data structure, which was saved and imported in JSON format.

### Rule-Based Inference Engine

We created a rule-based inference engine that can act as an expert system [9]. The system uses states and conditions organized in groups that change based on the input parameters and thereby evaluate clinical parameters against predefined logical rules. These rules determine which information entries are applicable to a given clinical scenario. The rule system was designed to handle:

- Simple equality conditions (e.g., tumor stage = pT1)
- Range conditions for numerical values (e.g., age < 40)
- Compound logical expressions using AND/OR/NOT operators
- Dependent conditions based on multiple parameters

Rules were formalized using a consistent syntax and stored in the database, allowing for maintenance and updates without modifying the application code. This separation of knowledge representation from application logic ensures that the system can evolve as clinical guidelines change.

Apart from these formal rules, we implemented a functionality for basic arithmetic calculations to compute additional relevant variables. Parameters and variables could further be re-used in rules and conditions.

### Web-Based User Interface

We implemented the user interface as a responsive web application using the Angular framework (version 15.2), providing cross-platform compatibility and accessibility through standard web browsers.

The interface architecture followed a component-based design pattern with three primary functional areas. The web-based user interface with an example input is shown in **Figure 2** and **Figure 3**. Additional Figures on the Parameter Input and SOP Results section, the Publications/Trials sand arithmetic functionalities are presented in the **Supplementary Figures**.

1. **Parameter Input Panel**: This panel contains all clinical parameter input controls organized by thematic categories (patient characteristics, tumor factors, treatment history). The input mechanisms were selected based on the data type of each parameter:

- Dropdown menus for categorical variables (e.g., tumor stage, molecular subtype)
- Numeric input fields as well as range sliders (which can be shown or hidden) for numerical variables (e.g., age, tumor size). The input of either exact values or ranges was possible (for cases of uncertainty/variance).
- Toggle switches for boolean parameters (e.g., presence of lymphovascular invasion)
2. **SOP Results**: The section below presents dynamically filtered knowledge base entries from the previous SOPs that match the current parameter configuration. Results are organized hierarchically by main topic and subtopic. Each entry provides:

- Concise clinical recommendation text
- Expandable rationale section explaining the recommendation logic
- Links to relevant other entries in the knowledge base or on the internet
- Visual indication of parameter dependencies affecting the recommendation
3. **Publications/Trials**: This section presents literature references relevant to the selected clinical scenario. The area is organized to present evidence-based medical knowledge.

- A list of clinical trials with visual indicators showing the strength of evidence.
- Publication listings with bibliographic information, including author, journal and year
- Interactive citation linking between SOP recommendations and their supporting evidence
- Further details with links to original sources on the internet provided after clicking on the individual entry

**Figure 2:**
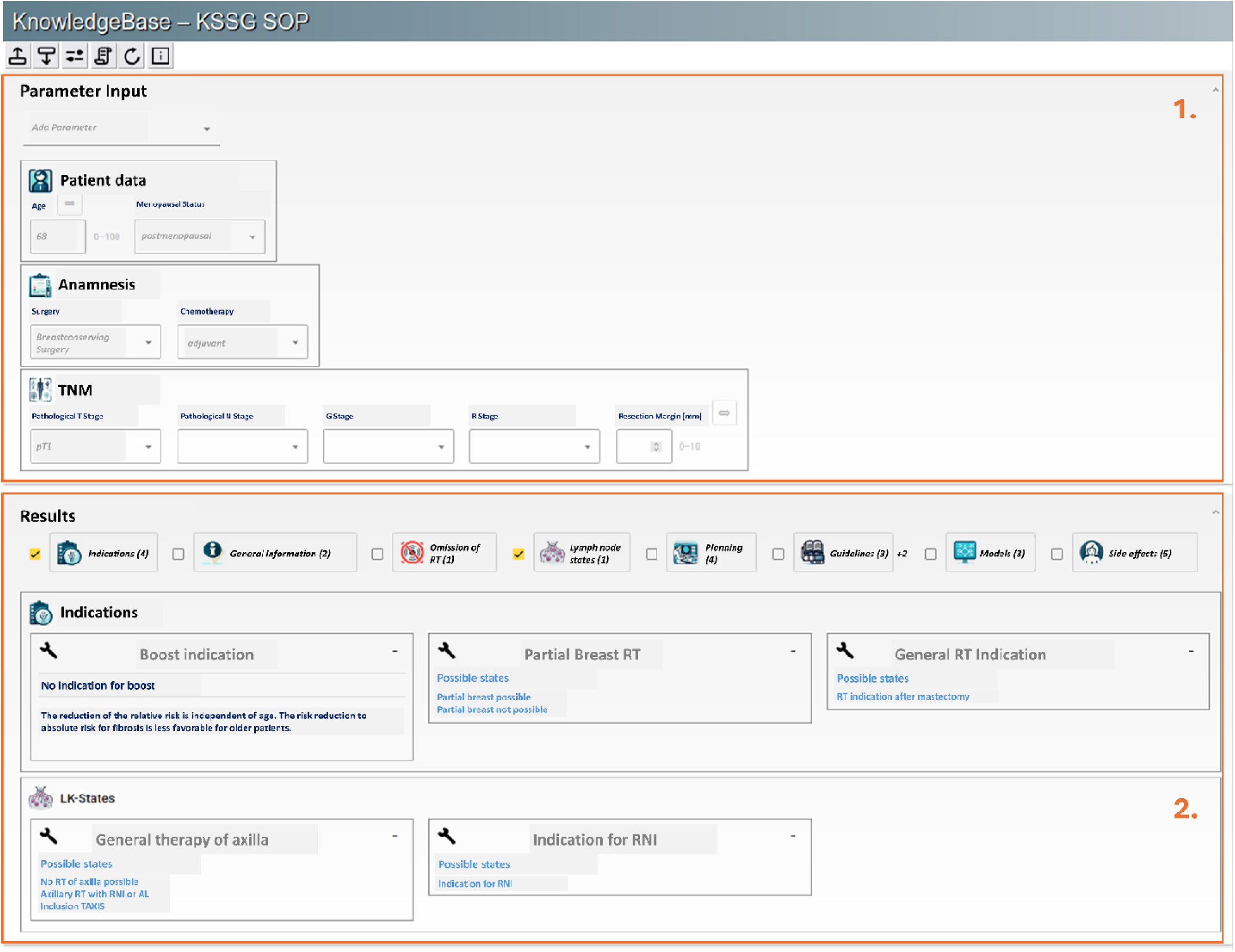
Upper part of our web-based user interface. It contains the Parameter Input Panel (1.) with the SOP Results section below (2.). Text translated in English for the Figure.

**Figure 3:**
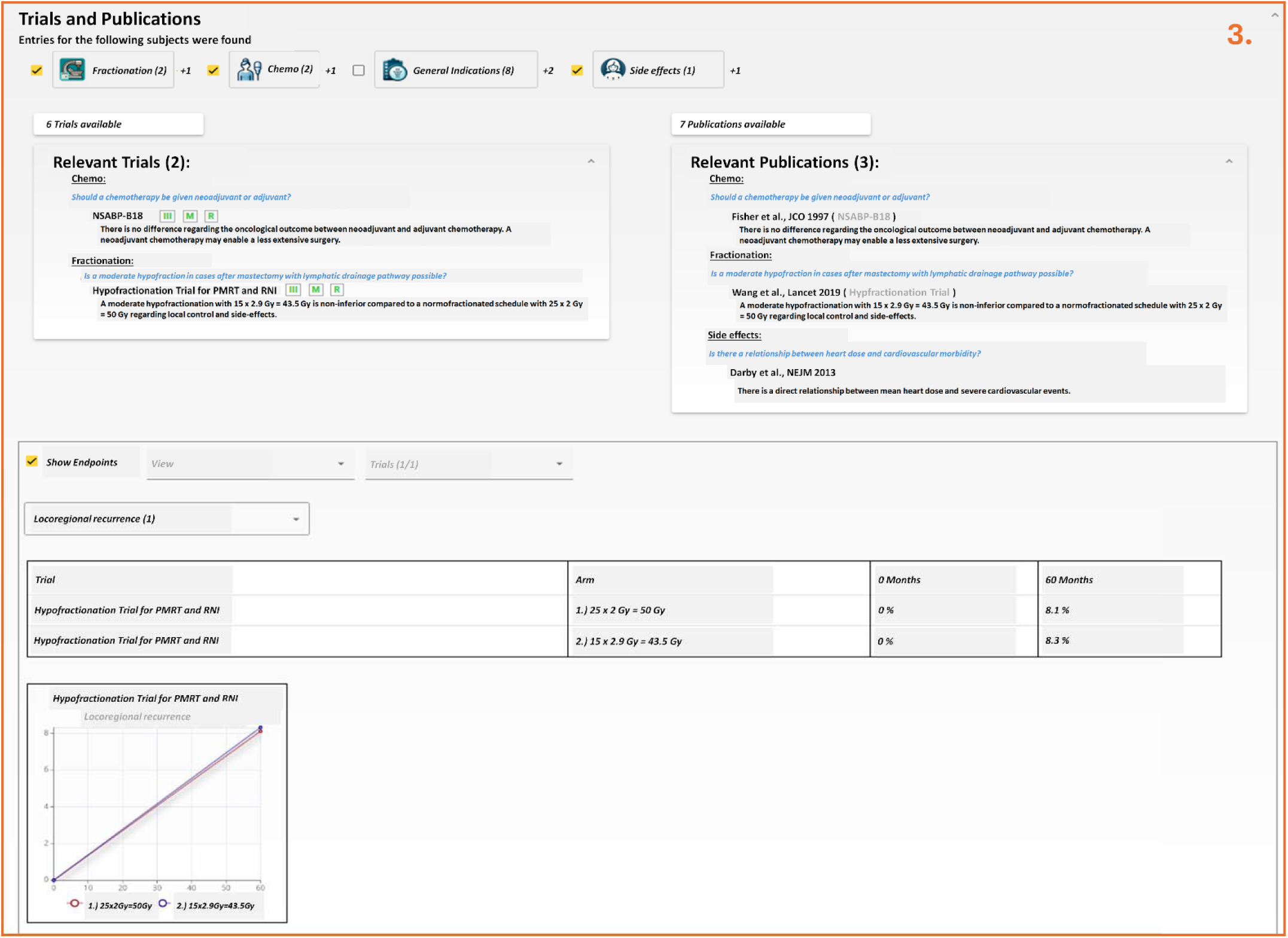
Lower part of our web-based user interface. It contains the Publications/Trials Results section. Text translated in English for the Figure.

### Testing

Testing was conducted through a structured evaluation process:

1. We recruited nine radiation oncologists from the Cantonal Hospital of St. Gallen for participating in the testing of the digital guidelines.
2. We collected usage data, including queries submitted, parameters specified, and usage time. Following data collection, the study coordinators manually classified the queries into appropriate query topics.
3. Through the web application, radiation oncologists could specify their information needs and provide feedback via an integrated form. They also indicated whether they found relevant information in the knowledge base. Success rates were measured by whether

users located pertinent information for their queries. For unsuccessful searches, we analyzed whether the requested information existed in the knowledge base. We evaluated user comments to determine if unsuccessful searches resulted from functional limitations or user experience issues.

The testing phase spanned over 14 months from 1^st^ of December 2023 to 31^st^ of January 2025.

## RESULTS

### Knowledge Base Content and Structure

The developed knowledge base incorporated comprehensive information on breast cancer radiotherapy structured in a hierarchical organization. The final system contained:

- 8 main topics covering different aspects related to radiotherapy management (indications, general information, omission of radiation therapy, lymph node status, radiation therapy planning, clinical guidelines, modeling and side effects).
- 31 subtopics providing more granular categorization of the knowledge domain
- 103 specific information entries containing detailed clinical guidance
- Information about 48 clinical trials relevant to breast cancer radiotherapy
- References to 65 scientific publications supporting the clinical recommendations

The content was written in German language. Lists of all input parameters, main topics and subtopics, are provided in **Appendix 1** and **Appendix 2**.

### Testing Results

At the end of the testing phase 62 distinct entries with complete data on query, parameters and evaluation were registered in the system. Most queries (33; 53.2%) were about “radiation therapy indication”. Further identified query topics included “Boost” (9; 14.1%), “treatment planning” (9; 14.1%), “lymph nodes” (8; 12.5%), “partial breast irradiation” (4; 6.3%) and “other” (1; 1.6%). Two of the queries were assigned to two topics.

Indication of successful information retrieval was indicated for 35 of the 62 queries (56.5%) while 27 queries (43.5%) were unsuccessful. Analysis of unsuccessful queries showed:

- For 21 of the 27 queries (77.8%), the requested information was available in the knowledge base, while in 6 cases (22.2%) it was not.
- For 19 of the unsuccessful queries, a free text comment by the user was provided, which indicated an issue regarding user experience in 15 cases (78.9%) and an issue in functional aspects in 4 cases (21.1%).

The majority of oncological scenarios used in the knowledge base were about early-stage breast cancer with 26 cases of staged tumor situations with pT1 and 31 cases with pN0.

## DISCUSSION

This study demonstrates the technical feasibility and clinical potential of a rule-based knowledge-base system for breast cancer radiotherapy. Our results highlight both additional capabilities of structured digital knowledge representation and the challenges of implementing such systems in clinical practice.

### Expert and Clinical Decision-Support Systems

The development of rule-based expert systems for clinical decision support has a rich history, dating back to early rule-based expert systems such as MYCIN [11], which demonstrated the potential for computational approaches to medical reasoning. While more modern systems increasingly used machine-learning (ML) algorithms, traditional rule-based approaches have several advantages. Most importantly, while ML-based systems function as “black boxes” [12], a rule-based approach provides transparency in the reasoning process, which is essential for clinician trust and adoption. This transparency allows users to understand why specific recommendations are presented, similar to the explainable approach advocated by Walsh et al. [13] in their review of decision support systems for oncology. Note that this is also beneficial from a legal perspective as it allows to justify decisions by rules and recommendations coming from SOPs and literature.

### Strengths and Innovations

Our knowledge base system demonstrates several notable strengths:

1. **Structured Knowledge Representation**: The hierarchical organization of radiotherapy knowledge into main topics, subtopics, and specific entries provides a logical framework that aligns with clinical thinking. This structure facilitates efficient navigation and retrieval of relevant information, addressing the fragmentation issues inherent in traditional text-based SOPs.
2. **Rule-Based Content Filtering**: The rule-based inference engine effectively transforms static guidelines into dynamic, context-sensitive recommendations. This approach has advantages over keyword-based searches by considering the logical relationships between clinical parameters.
3. **Evidence Integration**: The direct linking of recommendations to supporting clinical trials and publications enhances the evidence-based foundation of clinical decisions. This integration addresses a key limitation of traditional SOPs, which often become disconnected from their underlying evidence base as new research emerges.
4. **Modular Architecture**: The separation of knowledge content from application logic creates a sustainable platform that can evolve with changing clinical evidence without requiring re-development of the core system.

### Limitations and Challenges

Despite these conceptual strengths, our evaluation identified considerable limitations that must be addressed in future iterations. The clinical testing showed that a 43.5 % of queries were labeled unsuccessful by the users, while the detailed analysis indicated that data on most requests was actually available in the knowledge base. However, in most cases a query seemed to have been unsuccessful due to user-experience issues. The current version of the knowledge base is not adequately optimized for user-friendly interaction. The results demonstrate that emphasis on user experience is crucial to bring the extensive knowledge on complex topics like breast cancer to use. Technological sophistication must be balanced with simplicity in user interaction. As noted by Khairat et al. [14], even effective decision support systems fail if they create workflow disruptions or cognitive burdens.

Furthermore, despite the large amount of knowledge implemented, special scenarios are still not covered in the knowledge base. Complete coverage of all possible clinical scenarios is an asymptotic goal that requires ongoing knowledge curation.

The evaluation in the testing phase itself has some limitations. It focused primarily on technical functionality and immediate user feedback rather than long-term clinical impact. This leaves important questions about benefit unanswered. Furthermore, the initial testing with nine physicians from a single department and 62 queries is a limited number to draw conclusions from.

### Future Directions

Several technical considerations merit discussion and suggest directions for our future development:

1. **Content Maintenance Strategy**: The sustainability of any knowledge base depends on efficient processes for content updates as clinical evidence evolves. Automated approaches to evidence monitoring and rule generation could reduce the maintenance burden while improving currency.
2. **Expansion to Related Domains**: The CDE-based architecture developed for this breast cancer radiotherapy knowledge base could be extended to other cancer types or treatment modalities. Such expansion would benefit from the lessons learned in this implementation while broadening the system’s clinical utility.
3. **Potential for AI Enhancement**: While our current system relies on explicit rule formalization, future iterations could incorporate ML approaches. For example, ML might be used to extract structured data from free-text reports for improved data input.

Rule-based or “heuristic” expert systems have first been developed many decades ago and have demonstrated impressive success [11] [15]. Nevertheless, they failed to being broadly applied in medicine [16]. Reasons for this mainly include the immense complexity of medical knowledge [17], difficulty maintaining rules in rapidly evolving fields, lack of standardization across institutions [18], and limited ability to handle uncertainty and exceptions.

ML approaches subsequently gained prominence due to their ability to identify patterns in large datasets without explicit programming [19]. However, these systems often function as black boxes, creating challenges for clinical adoption where transparency and explainability are critical for trust and regulatory approval.

Rule-based systems may experience renewed relevance with the emergence of digital guidelines and interoperable data standards [20]. CDEs and standardized terminologies now provide the structured foundation these systems previously lacked. Furthermore, the increasing digitalization of clinical workflows creates natural integration points for rule-based decision support [21].

Furthermore, ML approaches and generative AI can be combined with explicit knowledge implemented in rule-based systems. This idea promises to overcome limitations of both approaches [22]. ML technologies can assist in automating rule curation, suggest updates based on new literature, translate guidelines into executable logic, and help maintain knowledge bases with reduced manual effort. At the same time humans can stay in the loop for oversight of rules implemented in a transparent way. Such a hybrid approach—combining the explainability of rules with the pattern-recognition capabilities of ML—may finally realize the clinical decision support potential envisioned decades ago.

## CONCLUSIONS

Using a rule-based knowledge base, we successfully digitalized SOPs on breast cancer radiotherapy into an interactive, evidence-linked resource using CDEs. Testing showed promising results, though substantial user-experiences barriers remain. Future development should prioritize interface refinement, sustainable content maintenance, and AI integration. By combining rule-based transparency with modern data standards and AI capabilities, interactive guidelines might be created that enhance evidence-based clinical practice while maintaining the explainability essential for medical applications.

## List of abbreviations

AI: Artificial Intelligence
CDE: Common Data Element
EHR: Electronic Health Record
FHIR: Fast Healthcare Interoperability Resources
ML: Machine Learning
NIH: National Institutes of Health
NPI: Nottingham Prognostic Index
SOP: Standard Operation Procedures

## Author contributions

Conceptualization – FD, JZ, NC, LP, PMP

Methodology, technical implementation – JZ, FD

Methodology, Testing – DW, MG, JC, KS, GFF, HLH, DB, LP

Statistical Analysis – FD

Writing, original draft preparation – FD

Writing, review and editing – All authors Project administration – FD, PMP

## Acknowledgments

Not applicable.

## Funding

The study received support through a grant from the Research Committee of HOCH Health Ostschweiz (Grant number 23/01).

## Disclosure of potential conflicts of interest

Dr. Cihoric is a technical lead for the *SmartOncology* project and medical advisor for Wemedoo AG, Steinhausen AG, Switzerland.

The authors declare no other conflicts of interest.

## Ethics approval

Not applicable. No patient-/person-related medical or non-medical data were used in this study. It therefore does not fall under the jurisdiction of the Federal Act on Research involving Human Beings (https:) and no approval from an ethics committee was required.

## Data and Code Availability Statement

This study did not involve any person-related data. The system is based on the local SOPs of the HOCH Cantonal Hospital St. Gallen, which belong to the institute and are not shared with the public. Details on the system architecture and the code are shared by the authors upon reasonable request. The source code of a related project on combining a knowledge base with a system for automatic data extraction in the form of CDEs is available under an MIT license at https://github.com/med-data-tools/data-extractor.

