## Appendix 1 for "Development of a Rule-Based Knowledge Base for Digitalization of Standard Operating Procedures on Radiotherapy in Breast Cancer"

**Appendix1: Details on Knowledge Base Content and Structure – Input Parameters**

The input parameters translated to CDEs are mostly identical to the ones published in our previous work on the creation of a CDE-based data structure for radiotherapeutic decision-making in breast cancer (<https://www.medrxiv.org/content/10.1101/2025.02.04.25321635v1.article-metrics>).

**Patient Data (Patientendaten)**

- **Age (Alter)**
  - Type: Number
  - Unit: year
  - Description: The current age (meaning at the time of the application of the data structure) of the patient in years.
- **Life expectancy (Lebenserwartung)**
  - Type: Number
  - Unit: year
  - Description: The estimated life expectancy of the patient in years.
- **Menopausal Status (prä-/postmenopausal):**
  - Type: Value List
  - Permissible Values: ‘premenopausal’ (prämenopausal), ‘postmenopausal’ (postmenopausal)
  - Description: Data on whether the patient is premenopausal or postmenopausal.

**Anamnesis (Anamnese)**

- **Type of oncological surgery conducted (OP):**
  - Type: Value List
  - Permissible Values: ‘breast conserving surgery’ (Brusterhaltende Operation), ‘mastectomy‘ (Mastektomie)
  - Description: Value about what general type (breast conserving or mastectomy) of oncological surgery has been conducted.
- **Chemotherapy conducted (Chemotherapie):**
  - Type: Value List
  - Permissible Values: ‘neoadjuvant’, ‘adjuvant‘, ‘none’ (keine)
  - Description: Value about the chemotherapy sequence (in case conducted).
- **Post-resection performed (Nachresektion erfolgt):**
  - Type: Value List
  - Permissible Values: ‘yes’ (ja), ‘no’ (nein)
  - Description: Binary value about whether a ‘post-resection’, meaning a second resection after first initial resection has been conducted.
- **Anti-Her2-Therapy ongoing/planned (Anti-Her2-Therapie):**
  - Type: Value List
  - Permissible Values: ‘yes’ (ja), ‘no’ (nein)
  - Description: Binary value about whether an Her2-targeted therapy is ongoing or planned.

**Further Details about the tumor (Weitere Angaben zum Tumor)**

- **extensive DCIS of the tumor lesion (Ausgedehntes intratumorales DCIS):**
  - Type: Value List
  - Permissible Values: ‘yes’, ‘no’
  - Description: Binary value about whether or not there is an extensive DCIS present.
- **Unifocal/multifocal (unifokal/multifokal):**
  - Type: Value List
  - Permissible Values: ‘unifocal’ (unifokal), ‘multifocal’ (multifokal)
  - Description: Value about whether there is an unifocal tumor lesion or there is a multifocal disease.
- **Laterality of tumor (Seite):**
  - Type: Value List
  - Permissible Values: ‘left’, ‘right’, ‘both’
  - Description: CDE about the laterality of the cancer disease.
- **Tumor location (Tumorlage):**
  - Type: Value List
  - Permissible Values: ‘lateral’, ‘medial’, ‘central’
  - Description: CDE about the location of the tumor lesions

**TNM:**

- **cT (klinisches T-Stadium):**
  - Type: Value List
  - Permissible Values: ‘cT1’, ‘cT2’, ‘cT3’, ‘cT4’
  - Description: CDE about the clinical T stage of the breast cancer situation (according to UICC).
- **cN (klinisches N-Stadium):**
  - Type: Value List
  - Permissible Values: ‘cN0’, ‘cN1’, ‘cN2’, ‘cN3’
  - Description: CDE about the clinical N stage of the breast cancer situation (according to UICC).
- **pT (pathologisches T-Stadium):**
  - Type: Value List
  - Permissible Values: ‘pT0’, ‘pTis’, ‘pT1’, ‘pT2’, ‘pT3’, ‘pT4’
  - Description: CDE about the pathological T stage of the breast cancer situation (according to UICC).
- **pN (pathologisches N-Stadium):**
  - Type: Value List
  - Permissible Values: ‘pN0’, ‘pN1’, ‘pN2’, ‘pN3’, ‘pN0(i+)’
  - Description: CDE about the pathological N stage of the breast cancer situation (according to UICC).
- **G status:**
  - Type: Value List
  - Permissible Values: ‘G1’, ‘G2’, ‘G3’
  - Description: Data about the grading (G status) of the cancer disease.
- **R status:**
  - Type: Value List
  - Permissible Values: ‘RX’, ‘R0’, ‘R1’, ‘R2’
  - Description: Value about the resection status (R status) of the cancer disease.
- **Minimal resection margin (Resektionsränder):**
  - Type: Number
  - Unit: mm
  - Description: Data value about the minimal resection margin of a tumor lesion

**Lymph node invastion (Lymphknotenbefall):**

- **Size of largest lymph node metastasis (Grösse LK-Metastase):**
  - Type: Number
  - Unit: mm
  - Description: The maximum size of the largest lymph node metastasis.
- **Number of positive sentinel lymph nodes (Anzahl befallener SN-LK):**
  - Type: Number
  - Unit: none
  - Description: The total number of positive sentinel (sn) lymph nodes.
- **Positive sentinel lymph node (SNL-Befall):**
  - Type: Value List
  - Permissible Values: ‘yes’ (ja), ‘no’ (nein)
  - Description: Binary value about whether there is a positive sentinel lymph node.
- **Number of positive resected lymph nodes (Anzahl befallene LK):**
  - Type: Number
  - Unit: none
  - Description: The total number of lymph nodes that were resected and had confirmed cancer of the breast cancer disease.
- **Number of totally resected lymph nodes (Anzahl resezierte LK):**
  - Type: Number
  - Unit: none
  - Description: The total number of lymph nodes that were resected as part of the breast cancer treatment, independent of whether or not they had confirmed cancer.
- **lymph node involvement in the mammaria interna region (Befall Mammaria-interna-LK):**
  - Type: Value List
  - Permissible Values: ‘yes’ (ja), ‘no’ (nein)
  - Description: Binary value about whether or not a lymph node in the mammaria interna region is positive.

**Receptors (Rezeptoren):**

- **Her2/neu-receptor status (Her2/neu-Rezeptorstatus):**
  - Type: Value List
  - Permissible Values: ‘positive’ (pos), ‘negative’ (neg)
  - Description: A binary value for the Her2/neu receptor status of the breast cancer disease (positive or negative). Even though it is recommended to use IHC class instead of positive/negative, the latter one is still used and can contain some information, which can be presented with this CDE.
- **Estrogen receptor status (ER):**
  - Type: Value List
  - Permissible Values: ‘positive’ (pos), ‘negative’ (neg)
  - Description: A binary value for the estrogen receptor status of the breast cancer disease (positive or negative).
- **Progesterone receptor status (PR):**
  - Type: Value List
  - Permissible Values: ‘positive’ (pos), ‘negative’ (neg)
  - Description: A binary value for the progesterone receptor status of the breast cancer disease (positive or negative).

**Further Pathological Markers (Weitere Pathomarker):**

- **LVI status:**
  - Type: Value List
  - Permissible Values: ‘positive’ (pos), ‘negative’ (neg)
  - Description: Binary value about the lymphovascular invasion (LVI status) of the cancer disease.
- **Histological subtype (Histologie):**
  - Type: Value List
  - Permissible Values: ‘no special type’ (NST), ‘invasive lobular’ (invasive-lobulär)
  - Description: Data about the histological subtype of breast cancer.
- **Tumor size (Tumorgrösse):**
  - Type: Number
  - Unit: mm
  - Description: Data value about largest diameter of the largest tumor lesion of a cancer disease.
- **Extracapsular extension of a lymph node metastasis (ECE):**
  - Type: Value List
  - Permissible Values: ‘yes’ (ja), ‘no’ (nein)
  - Description: Binary value about whether or not any positive lymph node had cancer with extracapsular extension.
- **Ki-67:**
  - Type: Number
  - Unit: %
  - Description: The %-value of the Ki-67 status of the breast cancer disease.

**Genetics (Genetik):**

- **BRCA1/2 status:**
  - Type: Value List
  - Permissible Values: ‘positive’ (pos), ‘negative’ (neg)
  - Description: Determines whether the patient has a mutation in the BRCA1/2 genes associated with a higher risk for breast cancer.
