## Appendix 2 for "Development of a Rule-Based Knowledge Base for Digitalization of Standard Operating Procedures on Radiotherapy in Breast Cancer"

**Appendix2: Details on Knowledge Base Content and Structure – SOP Results**

This list provides a basic overview of the topics and sub-topics implemented in the knowledge base.

**Indications (Indikationen)**

- Boost indications (Boostindikationen)
- Partial Breast RT (Teilbrust-RT)
- General RT Indications (Grundsätzliche RT-Indikation)
- Information about DCIS (Infos zu DCIS)

**General Information (Allgemeine Informationen)**

- Molecular Subtype (Molekularer Subtyp)
- (Internal States) (hidden)

**Omission of Radiation Therapy (RT-Verzicht)**

- Omission of Radiation (Verzicht auf Bestrahlung)

**Lymph node status (LK-States)**

- General Therapy of the Axilla (Allgemeine Therapie der Axilla)
- Indication for regional nodal irradiation (Indikation für RNI)

**Radiation Therapy Planning (Planung)**

- Fractionation (Fraktionierung)
- Planning CT (Planungs-CT)
- Contouring (Konturierung)
- Dose constraints

**Guidelines**

- **Guidelines / ASTRO**
  - ASTRO Partial Breast Guideline
  - ASTRO Boost Recommendation
- **Guidelines / AGO**
  - AGO-Guideline

**Modelling (Modelle)**

- NPI-Values
- NPI N (hidden)
- NPI G (hidden)
- Predict
- CTS5 Calculator
- MSKCC Nomogram – Sentinel Lymph Node Metastasis
- MSKCC Nomogram – Additional Non SLN Metastases
- MSKCC Nomogram – DCIS Recurrence
- KCI – Overall Survival Curve
- KCI – LRR in Postmastectomy N0

**Side effects (Nebenwirkungen)**

- Radiodermatitis
- Nausea (Übelkeit)
- Pruritus
- Fear (Angst)
- Pain (Schmerzen)
