## Supplementary Figures for "Development of a Rule-Based Knowledge Base for Digitalization of Standard Operating Procedures on Radiotherapy in Breast Cancer"

**Supplementary Figure 1** shows how an example oncological scenario and different parts of the user interface including the parameter input section (**Supplementary Figure 1A**), the results display of an SOP recommendation (**Supplementary Figure 1B**) as well as details on the recommendations and rules for the entry implemented in the knowledge base (**Supplementary Figure 1C**).

**Supplementary Figure 2** shows how in a similar way the entries for the Publications/Trials section were provided based on the input parameters.

**Supplementary Figure 3** demonstrates how the arithmetic functionalities were used to calculate prognostic scores and provide additional relevant information to the user.

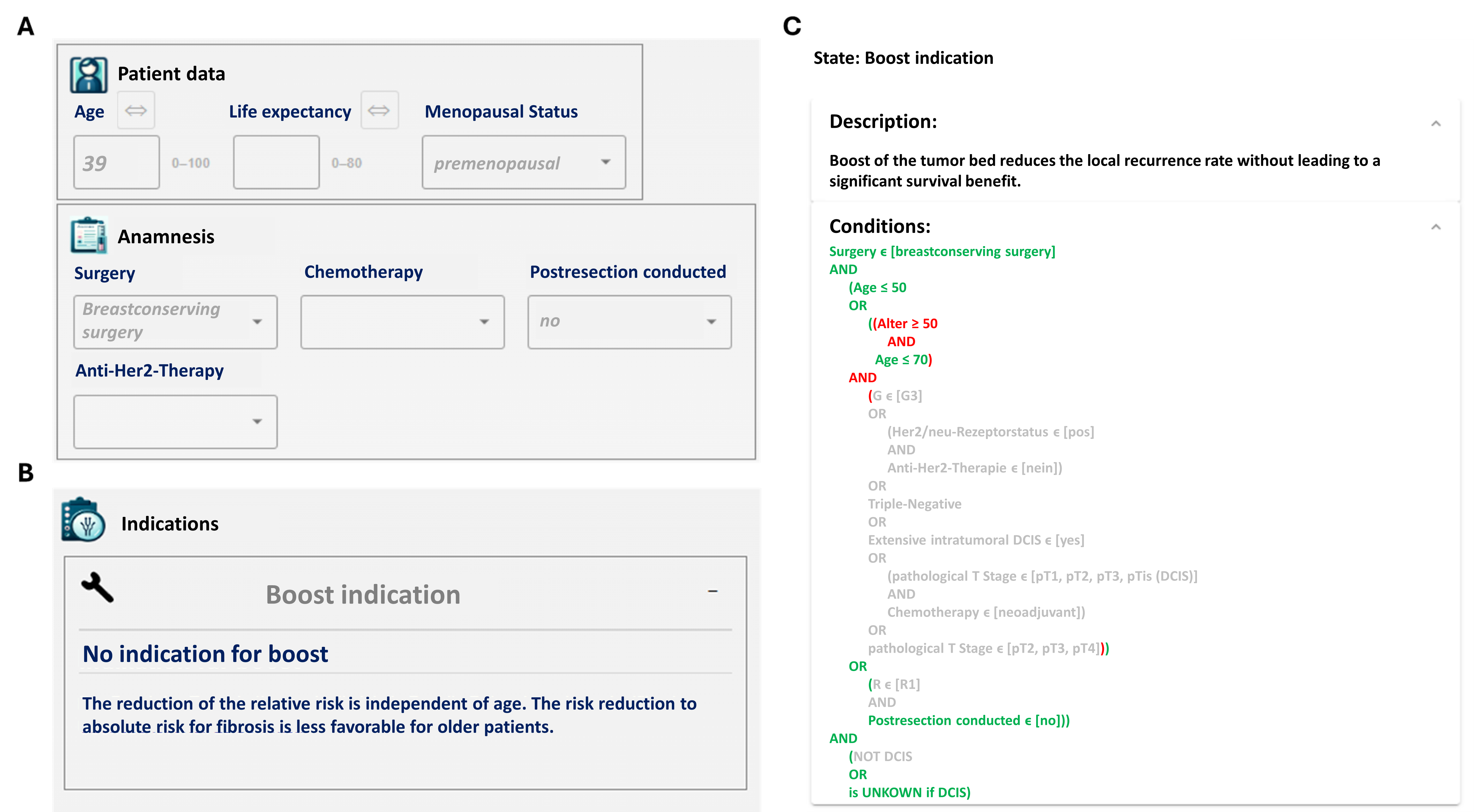

Supplementary Figure 1: Different parts of the Parameter Input and SOP Results section of the rule-based knowledge base user interface.
A – Input panel for categorical and numerical values organized into different sections (like “patient data” or “anamnesis”).
B – SOP Result section showing relevant entries based on the input parameters (example: indication for boost irradiation).
C – Details on the obtained result with further description as well as the rules defined in the knowledge base for the entry. Text translated in English for the Figure.

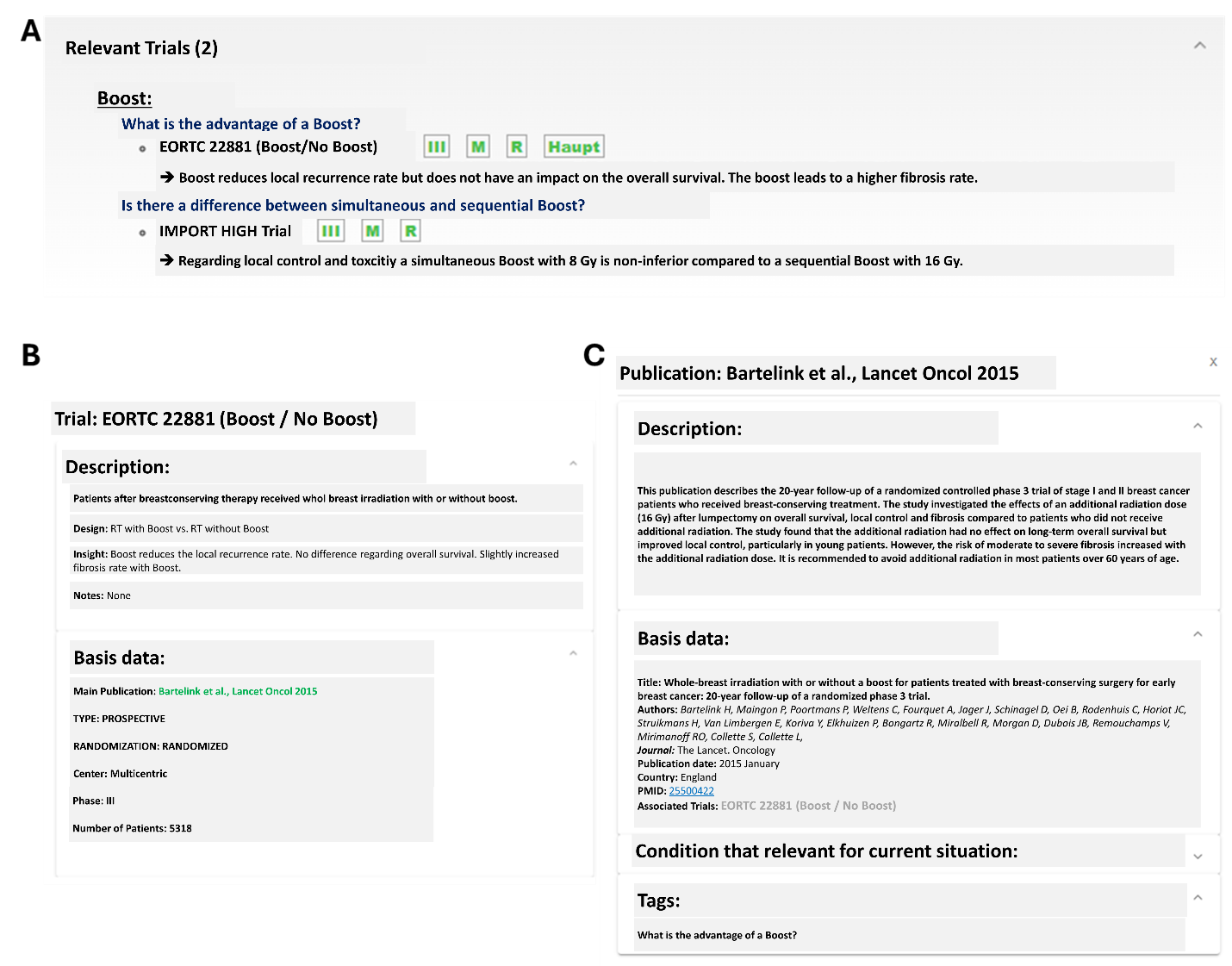

Supplementary Figure 2: Different parts of the Publications/Trials section of the rule-based knowledge base user interface.
A – Relevant trials shown to given input parameters with visual indicators for evidence overview (III – Phase 3 Trial, M – Multicentric Trial, R – Randomized Trial, Haupt – Main criteria that were required for inclusion in this trial are fulfilled).
B – Dialog with additional details on a selected trial. Note that this entry includes a link to the related publication.
C – Dialog with details on a selected publication; it includes a link to the original source in a database such as PubMed. Text translated in English for the Figure.

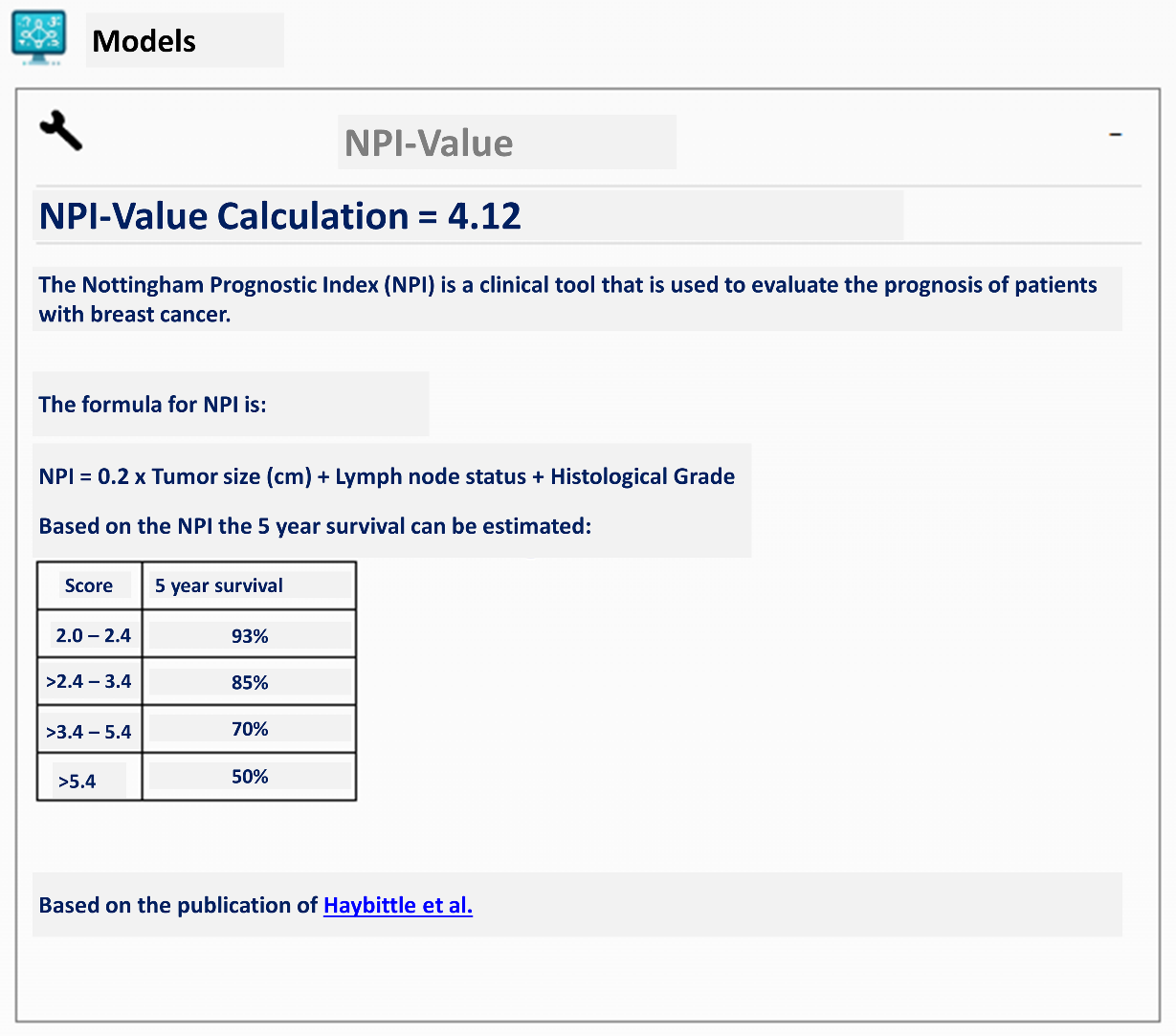

Supplementary Figure 3: Entry making use of the arithmetic functionalities implemented in the system. In this example, the Nottingham Prognostic Index (NPI) is directly calculated based on the input parameters tumor size, lymph node size, and histological grade based on the work of Haybittle et al. [10]. Based on the NPI, the prognosis of the patient can be estimated. Text translated in English for the Figure.
